# Gaps in protection to *Anopheles* exposure in high malaria endemic regencies of Papua Province, Indonesia

**DOI:** 10.1101/2024.09.17.24313788

**Authors:** Ismail E Rozi, Lepa Syahrani, Dendi H Permana, Puji B S Asih, Maria E Sumiwi, Neil F Lobo, William A Hawley, Din Syafruddin

**Affiliations:** Eijkman Research Center for Molecular Biology, National Research and Innovation Agency (BRIN), Cibinong, Indonesia; Doctoral Program in Faculty of Medicine, Hasanuddin University, Makassar, Indonesia; Doctoral Program in Faculty of Mathematics and Natural Sciences, University of Indonesia, Depok, Indonesia; Doctoral Program in Faculty of Medicine, University of Indonesia, Jakarta, Indonesia; United Nations Children’s Fund (UNICEF), Jakarta, Indonesia; Eck Institute for Global Health, University of Notre Dame, Indiana, USA; Department of Parasitology, Faculty of Medicine, Hasanuddin University, Makassar, Indonesia; Hasanuddin University Medical Research Center, Makassar, Indonesia

**Author notes:** Correspondent author Hasanuddin University Medical Research Center, Makassar, Indonesia.

**Keywords:** Human behavioural observation, gaps in protection against mosquito bite, malaria, Papua Province, Indonesia

## Abstract

**Background:** Malaria in eastern Indonesia remains high despite significant reductions and local elimination in other parts of the country. Malaria control activities that have been implemented include early diagnosis and prompt treatment, provision of Long-lasting insecticide-treated nets (LLINs), and indoor residual spraying (IRS). To expedite malaria elimination in this region, a rapid entomological assessment combined with human behaviour observations (HBOs) were conducted in eight high malaria endemic regencies in Papua Province, Indonesia. The present study focuses on identifying gaps in protection against mosquito biting indoors and outdoors that may contribute to the sustained transmission and persistently high endemicity.

**Methods:** This study was conducted alongside a rapid entomologic assessment, including human landing catches (HLCs) of adult mosquitoes. Human behavior was documented by direct observations during HLCs. HBO data focused on temporal (over the night) and spatial (domestic or peri-domestic) presence, alongside bed net usage and sleeping patterns. Household questionnaires, also conducted during entomological collections, documented data on house structure materials, practices against mosquito bites, livestock presence, as well as intervention usage.

**Results:** Analysis of human behaviors in each regency identified several indoor and outdoor gaps in protection against mosquito biting. Human exposure to mosquito bites was driven by ITN usage, IRS coverage, indoor presence without protection prior to sleeping, absence of mosquito house screens, and outdoor presence without protection.

**Conclusions:** Data demonstrates multiple gaps in protection against mosquito exposure in all eight regencies of Papua evaluated. Indoor interventions require optimization, while current vector control activities do not presently address outdoor exposure. Measured spatial and temporal exposure may be utilized to understand protective vector interventions that may function in these spaces while also pointing to continued exposure. Additional interventions, such as community-based larval source management, may reduce exposure overall, while novel interventions like spatial repellents may fill some gaps in protection – alongside optimized case detection and treatment. Results suggest that the present strategy may be insufficient to eliminate malaria in the region, and a rethought evidence-based and adaptive strategy is required.

## INTRODUCTION

Malaria incidence in Indonesia has been successfully brought down in many parts of the country except for some Provinces in the east, namely Papua, West Papua, and East Nusa Tenggara [1]. Of the 443,530 Indonesia malaria cases reported in 2022, 393,801 cases, or more than 88%, were from Papua. There are eight regencies in Papua Province, all of which have an Annual Parasite Incidence (API) of more than 100 [1–3]. In addition, the COVID-19 pandemic in 2019 negatively affected healthcare services, including malaria control activities, at the primary health center level across Indonesia [4], resulting in concerns of increased incidence, outbreaks as well as reintroduction of malaria in areas where it had been eliminated.

As a response, the government of Indonesia has, within the last few years, intensified malaria control activities in Papua. These enhanced activities include better malaria case detection and treatment based on reviving village malaria cadres that enable early detection and prompt treatment of cases [5,6]. Vector interventions that supplement these epidemiological strategies include the targeted full coverage with both Insecticide-treated nets (ITNs) and indoor residual spraying (IRS) [7] in malaria-endemic regencies.

Indonesia has set an ambitious goal to eliminate malaria by 2030 [8], targeting eradication district by district. As of 2022, 72.37% of districts (equivalent to 372 districts) across the country have been declared free of malaria transmission [1,2]. However, none of the districts in Papua have achieved this status. Papua faces the most significant challenge in meeting the 2030 malaria elimination target due to its current high endemicity. In 2023, eight districts in Papua—Jayapura, Yapen Islands, Mimika, Boven Digoel, Asmat, Sarmi, Keerom, and Waropen—were reported to have high malaria incidence [3]. These regencies have implemented the three pillars of malaria control: early diagnosis and prompt treatment, distribution of ITNs, and IRS. Despite these efforts, the annual malaria parasite incidence has remained high, seen even before the COVID-19 pandemic.

To accelerate the reduction of malaria cases and achieve the 2030 elimination goal, rapid entomological assessments, household surveys, and observations of human behavior toward understanding the transmission system were conducted in five districts in 2019 and three more in 2021. "Gaps in protection" are situations where individuals or households are potentially exposed to malaria due to the absence or inadequacy of effective protective or preventive measures [9,10]. These gaps can be identified by assessing how interventions interact with local human and vector populations. In this study, identifying and characterizing these gaps in protection includes examining the presence and use of ITN, IRS coverage, housing conditions, and spatial and temporal human activities and presence [10]. By analyzing data from human behavior and household surveys in these eight districts, the study aims to pinpoint the gaps in protection against mosquito bites, indoors and outdoors, that may contribute to the persistently high malaria cases in Papua.

## METHODS

### Ethical statement

This study was approved by the Ethics Committee of Research in Health, Medical Faculty of Hasanuddin University, Makassar, Indonesia No: 281/UN4.6.4.5.31/PP36/2019 and (29 January 2021) No: 40/UN4.6.4.5.31/PP36/2021

### Site selection

This study was conducted as part of the Rapid Entomological Assessments program across eight regencies in Papua province, Indonesia (Fig 1). In each regency, three health centers were selected, and within each health center’s jurisdiction, two villages were chosen for the study. A total of 48 villages were included, comprising 10 transmigrant communities and 38 resident settlements. The entomological assessments and household surveys were carried out across all eight regencies over 150 days.

**Fig 1.**
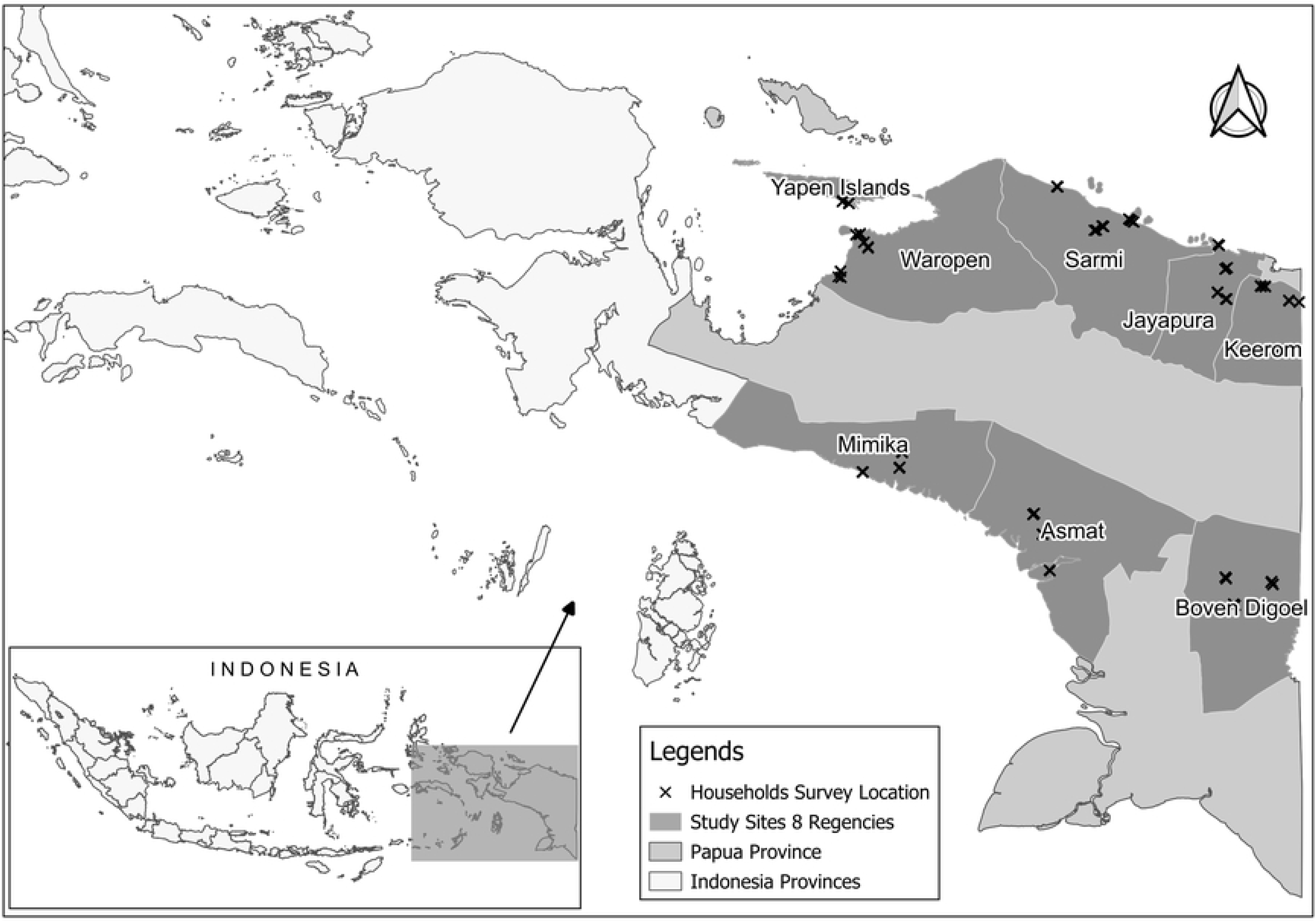
Location of study sites. Location of the study sites in eight regencies of Papua Province: Keerom, Jayapura, Sarmi, Mimika, Boven Digoel, Yapen Islands, Waropen, and Asmat

### Adult mosquito collections

Adult mosquito collections were performed using HLCs and indoor night resting collection [11]. HLCs were conducted in four sentinel houses in each area and performed from 1800 to 0600 to evaluate *Anopheles*’ indoor and outdoor landing densities in the houses. Human biting rate (HBR) indoors and outdoors were calculated as bite per person per hour (bph). The indoor and outdoor HBRs were used to calculate the periodicity of *Anopheles* biting activities per site and utilized to estimate human behavior-adjusted biting rates [12].

Meanwhile, indoor night resting collections were performed using manual aspiration every two hours from 1800 to 0600 in the four houses without HLC activities per site. The indoor resting density (IRD) was calculated as the total *Anopheles* collected per sentinel house per night. The estimated IRS effectiveness for potential protection in each study site, when applied, was calculated by comparing the total *Anopheles* captured per house from HLC divided by IRD [13]. This calculation was also carried out for each species of Punctulatus group collected. HLCs and indoor night resting collections were conducted by trained local human volunteers, along with signed informed consent regarding the possible effects and risks of participating in the study [12,14].

### Human behavior observations (HBOs)

HBOs were conducted at the same time as HLCs. At the end of every HLC hour, the HLC volunteer documented the number of people indoors and whether they were using an ITN. Similarly, the outdoor HLC volunteer documented the number of people within a 5 m radius of the house (the peridomestic area) with the same spatial and temporal behavioral endpoints (S1 File Part A). Analysis to determine human behavior-adjusted exposure was based on the overlap between spatial and temporal presence alongside ITN use and parallel vector data [15–18]. The proportions of HBOs were calculated separately using 48 HBOs data of each study site location. The potential risk of exposure to *Anopheles* vectors was only calculated in locations where a high number of *Anopheles* species were found.

### Household survey

A community-based household survey was conducted with the assistance of local health workers during household enrollment near mosquito breeding site sampling locations. The survey aimed to enroll 20 to 30 households per village across 48 villages. The survey gathered information on building materials, socio-economic status, nighttime human behavior, protection against mosquito bites, and the presence of malaria cases in the households (refer to S1 File Part B). Data collection was facilitated using ODK Collect, a mobile platform that operates on the Open Data Kit (ODK) system [19]. Quantitative data was analyzed using basic functions in Microsoft Office Excel and open-source software, RStudio version 1.3.1056, based on R version 4.0.2 [20,21]. The k-means algorithm from the klaR package was employed to cluster house types based on material structures [22]. The analysis primarily involved calculating percentages and proportions. ITN access was determined by dividing the total number of potential ITN users by the total number of individuals who spent the previous night in the surveyed households [23–25].

To assess the socioeconomic status of the households in the study area, a wealth index (WI) was calculated using a principal component analysis (PCA) in R [20,21]. This approach was based on the methods used in demographic and health surveys (DHS) for wealth index calculations [26–28]. A total of 23 key asset ownership variables, including house amenities and household productive and non-productive assets, were collected from the household survey and combined into proxy WI indicators using a PCA. The resulting wealth index was then used to rank households into five quintiles, which were prepared for cross-tabulation with other variables in the dataset.

### Gaps in protection

Gaps in protection [9]were represented by HBO-adjusted biting rate, i.e. exposure to mosquito bites, house structures including materials and protection from mosquitoes entering the house (installment of window screens and the use of indoor repellents), the distribution and use of ITNs, the coverage of IRS, and individual human night time behaviors related to exposure to mosquito bites including sleeping behaviors [9,10]. The magnitude of gaps in protection was indicated by symbols "+," "++," and "+++", representing minor, moderate, and major issues, respectively. These were calculated based on the proportion of related circumstances within the entire population. A chi-square test was employed to determine whether the gaps in protection were statistically significantly associated (using a critical p-value of 0.05) with clustered house types and the socioeconomic status of households in the study area.

## RESULTS

### Human behavior observations during HLC

The presence and activities of all inhabitants of the sentinel houses during the HLC period is shown in Fig 2a. Human activities were divided into four behavioral groups: indoor under ITNs, indoor not under ITNs, outdoor-awake, and outdoor-sleep. Both outdoor behavioral groups did not use ITNs. Many data points were missed during the HBOs, particularly in the early hours (18:00-19:00) and the final periods (04:00-06:00) of the HLC observations. Moreover, additional missing data was identified from five villages in Yapen Islands Regency, which were represented as zero values in stacked area charts. HBOs conducted between 19:00 and 20:00 indicated limited outdoor activity, with over 70% of observed activities occurring indoors. More than 65% of the people indoors were not under ITNs. Gradually, the proportion of individuals using ITNs increased, peaking after midnight. From midnight to 06:00, the percentage of people indoors remained steady between 30% and 40%, suggesting that most slept indoors without ITNs. Since the proportion of individuals sleeping outdoors was minimal, it was combined with the proportion of those awake outdoors in subsequent analyses, referred to as the proportion of humans outdoors.

**Fig 2a.**
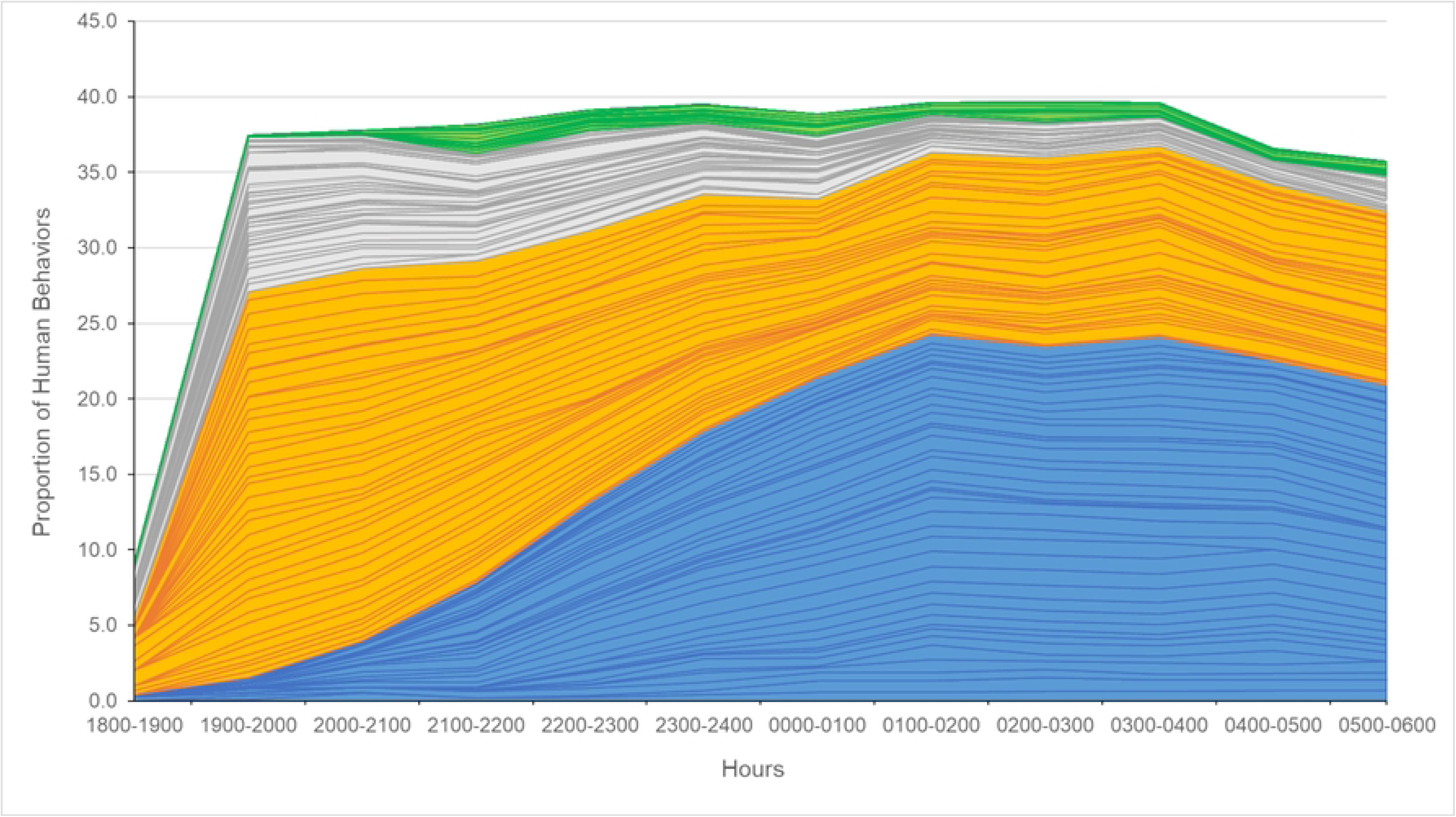
Human behavior proportions from all study sites. Stacked area charts in blue showed proportions from indoor under ITNs, orange for indoor not under ITNs, grey for outdoor-awake dan green for outdoor-sleep.

Fig 2b illustrates box-and-whisker plots showing the total proportions of human behavior across all sites and within each regency. Data from the Yapen Islands was excluded, and the categories "outdoor awake" and "outdoor sleeping" were combined into a single "outdoors" category. The HBO results showed that the proportions of people indoors under ITNs, indoors without ITNs, and outdoors across all regencies were 38.2%, 47.7%, and 14.1%, respectively. Among the regencies, Sarmi had the highest percentage of people indoors under ITNs and the lowest percentage indoors without ITNs. In contrast, the overall outdoor population was low, with Waropen and Asmat regencies recording the smallest outdoor populations.

**Fig 2b.**
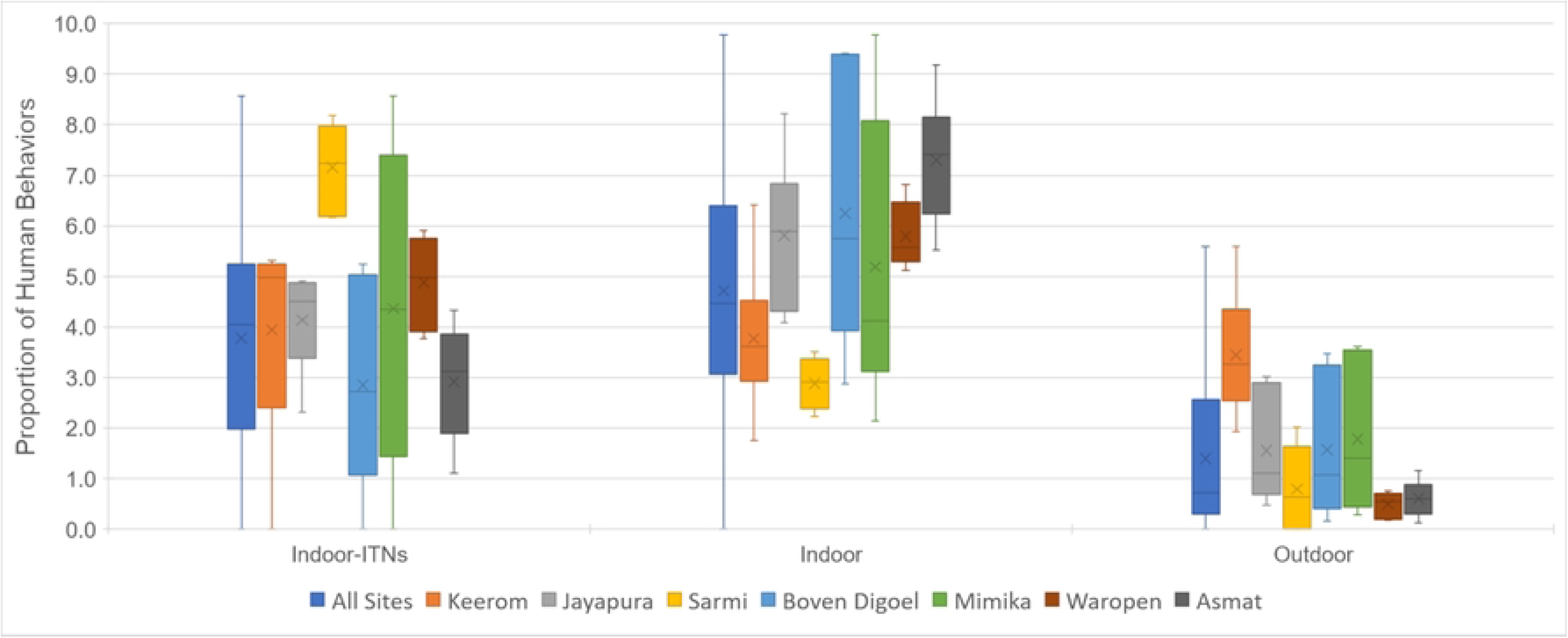
Total proportions of human behavior for each regency.

### Human behavior adjusted mosquito biting rate

S2 File presents the total proportion of human behavior, the proportion of human behavior overlaid with indoor and outdoor mosquito biting rates, and the human behavior-adjusted biting rate for an unprotected individual’s exposure to *Anopheles* bites. This data is derived from the 12 villages with high human biting rates per night from HLC, including nine villages from inland areas and three from coastal regions. Additionally, data from two more villages—Migiwia, a coastal village in Mimika, and Yasiuw, a riverside village in Asmat—are also included.

Table 1 displays the directly measured mosquito biting as well as the proportions of behavior-adjusted exposure for ITN users and unprotected individuals in these 14 villages. Assuming that individuals under ITNs are fully protected from *Anopheles* bites, villages with a high proportion of the population using ITNs during peak biting times would experience greater protection (e.g., villages in Sarmi Regency such as Samanente, Konderjan, and Webrau, with 74.4%, 68.5%, and 68.2% of the population under ITNs, respectively).

**Table 1.**
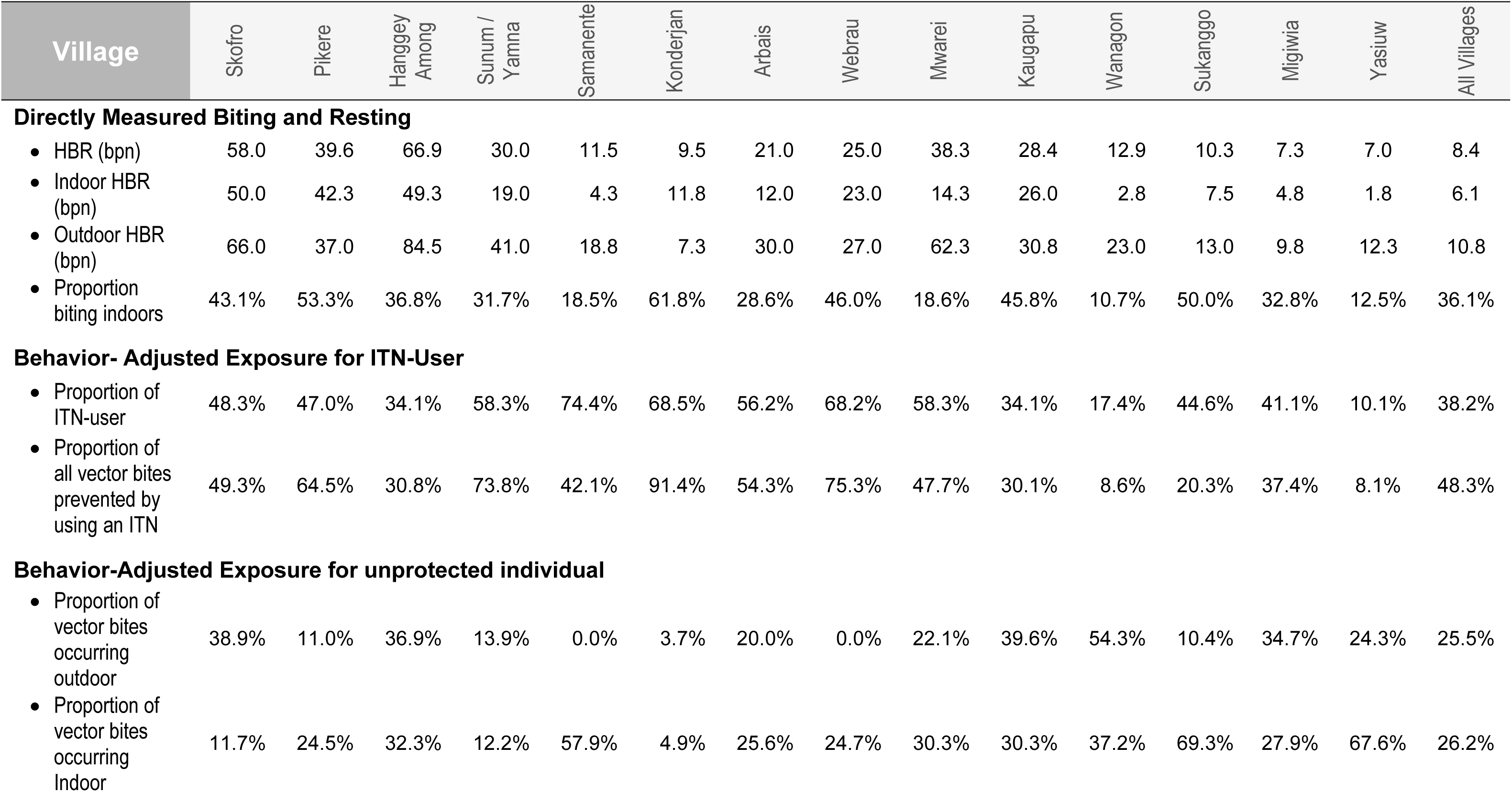
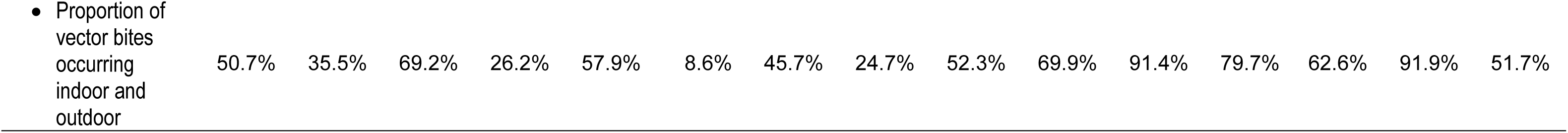
Behavior-adjusted *Anopheles* exposure of 14 villages.

| Village | Skofro | Pikere | Hanggey<br>Among | Sunum /<br>Yamna | Samanente | Konderjan | Arbais | Webrau | Mwarei | Kaugapu | Wanagon | Sukanggo | Migiwia | Yasiuw | All Villages |
| --- | --- | --- | --- | --- | --- | --- | --- | --- | --- | --- | --- | --- | --- | --- | --- |
| **Directly Measured Biting and Resting** |  |  |  |  |  |  |  |  |  |  |  |  |  |  |  |
| • HBR (bpn) | 58.0 | 39.6 | 66.9 | 30.0 | 11.5 | 9.5 | 21.0 | 25.0 | 38.3 | 28.4 | 12.9 | 10.3 | 7.3 | 7.0 | 8.4 |
| • Indoor HBR (bpn) | 50.0 | 42.3 | 49.3 | 19.0 | 4.3 | 11.8 | 12.0 | 23.0 | 14.3 | 26.0 | 2.8 | 7.5 | 4.8 | 1.8 | 6.1 |
| • Outdoor HBR (bpn) | 66.0 | 37.0 | 84.5 | 41.0 | 18.8 | 7.3 | 30.0 | 27.0 | 62.3 | 30.8 | 23.0 | 13.0 | 9.8 | 12.3 | 10.8 |
| • Proportion biting indoors | 43.1% | 53.3% | 36.8% | 31.7% | 18.5% | 61.8% | 28.6% | 46.0% | 18.6% | 45.8% | 10.7% | 50.0% | 32.8% | 12.5% | 36.1% |
| **Behavior- Adjusted Exposure for ITN-User** |  |  |  |  |  |  |  |  |  |  |  |  |  |  |  |
| • Proportion of ITN-user | 48.3% | 47.0% | 34.1% | 58.3% | 74.4% | 68.5% | 56.2% | 68.2% | 58.3% | 34.1% | 17.4% | 44.6% | 41.1% | 10.1% | 38.2% |
| • Proportion of all vector bites prevented by using an ITN | 49.3% | 64.5% | 30.8% | 73.8% | 42.1% | 91.4% | 54.3% | 75.3% | 47.7% | 30.1% | 8.6% | 20.3% | 37.4% | 8.1% | 48.3% |
| **Behavior-Adjusted Exposure for unprotected individual** |  |  |  |  |  |  |  |  |  |  |  |  |  |  |  |
| • Proportion of vector bites occurring outdoor | 38.9% | 11.0% | 36.9% | 13.9% | 0.0% | 3.7% | 20.0% | 0.0% | 22.1% | 39.6% | 54.3% | 10.4% | 34.7% | 24.3% | 25.5% |
| • Proportion of vector bites occurring Indoor | 11.7% | 24.5% | 32.3% | 12.2% | 57.9% | 4.9% | 25.6% | 24.7% | 30.3% | 30.3% | 37.2% | 69.3% | 27.9% | 67.6% | 26.2% |

- Proportion of vector bites occurring indoor and outdoor
|  |  |  |  |  |  |  |  |  |  |  |  |  |  |  |
| --- | --- | --- | --- | --- | --- | --- | --- | --- | --- | --- | --- | --- | --- | --- |
| 50.7% | 35.5% | 69.2% | 26.2% | 57.9% | 8.6% | 45.7% | 24.7% | 52.3% | 69.9% | 91.4% | 79.7% | 62.6% | 91.9% | 51.7% |

The behavior-adjusted exposure rate for unprotected individuals revealed different characteristics across the villages, influenced not only by observed human behavior but also by geographical factors and the predominant *Anopheles* species’ biting behavior in the area. In most of the 12 villages, *An. koliensis* was the dominant species, with peak biting occurring after midnight. However, in two inland villages in Mimika Regency - Mwarei and Kaugapu - *An. farauti* was found in significant numbers during the early evening, with fewer *An. koliensis* collected at that time (Figs S2-9b and S2-10b). The presence of this species and their biting behaviors increased the risk of exposure to *Anopheles* bites for populations not yet under ITNs. The proportions of vector bites occurring indoors and outdoors in Mwarei and Kaugapu were 52.3% and 69.9%, respectively. Minimal outdoor activity after 18:00-19:00 in Mwarei resulted in a relatively low adjusted exposure rate for unprotected individuals (Fig S2-9d). This pattern was also observed in other sites where *Anopheles* began seeking blood meals indoors early in the evening.

In Migiwia, a coastal village in Mimika Regency, *An. farauti* was the predominant species, with blood-seeking activity peaking in the early evening, shortly after midnight, and again toward the end of the night (HBR indoors and outdoors was 4.8 bpn and 9.8 bpn, respectively; Fig S2-13b). In Yasiuw, a riverside village in Asmat Regency, high blood-seeking activity early in the night was primarily due to *An. koliensis* (HBR indoors and outdoors were 1.8 bpn and 12.3 bpn, respectively; Fig S2-14b).

Overall, in most villages, there was a high risk of exposure to *Anopheles* bites before midnight, when most of the population was not yet under ITNs. In villages with extremely high HBR, such as Skofro and Hanggey Among, even though a large portion of the population was under ITNs during observation, the remaining indoor and outdoor populations were still highly exposed to *Anopheles* bites throughout the night, with proportions of vector bites occurring indoors and outdoors at 50.7% and 69.2%, respectively (Figs S2-1d and S2-3d). Moreover, villages with a lower proportion of the population under ITNs, such as Wanagon, Sukanggo, and Yasiuw, were also highly exposed to *Anopheles* bites (Figs S2-11d, S2-12d, and S2-14d), with proportions of vector bites occurring indoors and outdoors at 91.4%, 79.7%, and 91.9%, respectively.

### Demographic characteristics

The results of the household questionnaire, conducted across 1,255 households in 47 villages, are presented in Supplementary File 3, Tables S3-1 to S3-5. Data from one village in Yapen Islands Regency was missing due to the loss of the mobile device used for data collection before it could be uploaded. The gender ratio of interviewees across the eight regencies was not significantly different, with 86.2% of respondents being the head of the household or their spouse (Table S3-1). The remaining 10.8% were primarily other primary family members, such as children, grandchildren, parents, or parents-in-law. The average age of respondents was 40.7 years (±14.1), with ages ranging from 12 to 83 years. Regarding education levels, 39.8% of respondents had completed senior high school or higher, 47.0% had finished primary or junior high school, and the remainder either had no formal education or did not complete elementary school.

The average number of households surveyed per village was 26.7 (±4.7) (Table S3-2), with an average household size of 5.6 (±3.5) members. The majority of surveyed households fell into the lower socioeconomic categories, primarily in the second and first quantiles, classified as poorer quantiles. Regarding housing structure, 61% of the homes in the eight regencies were constructed from wooden planks (with full or half-walls made of wooden planks) and had zinc roofs (Figs 3a, 3c, and 3d). Another 37% of the houses were built with brick, cement, or stone walls (Fig 3b). Almost 93% of the houses in the study areas had zinc roofs, while the remaining roofs were made from asbestos, ceramic tiles, or thatch.

**Fig 3.**
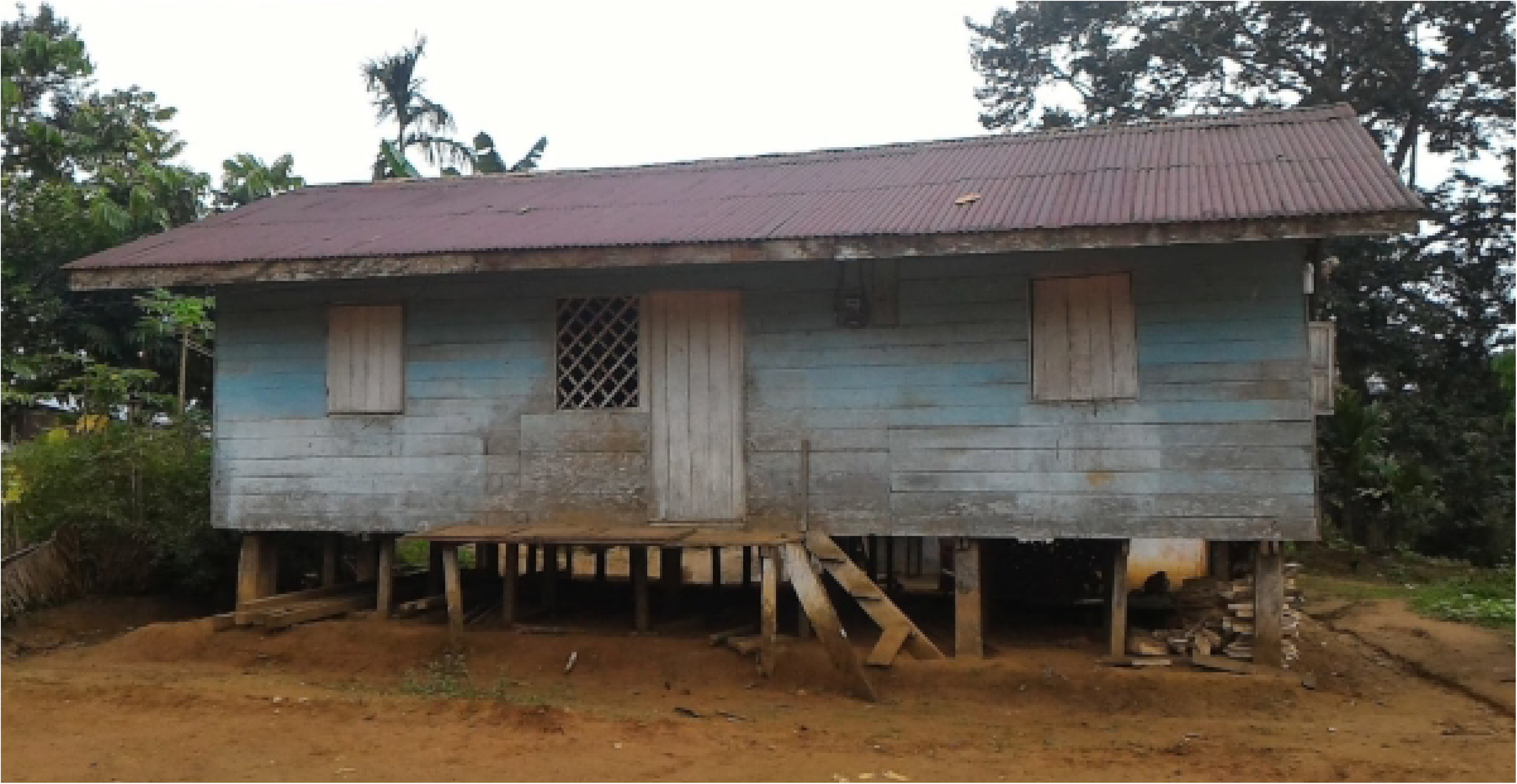
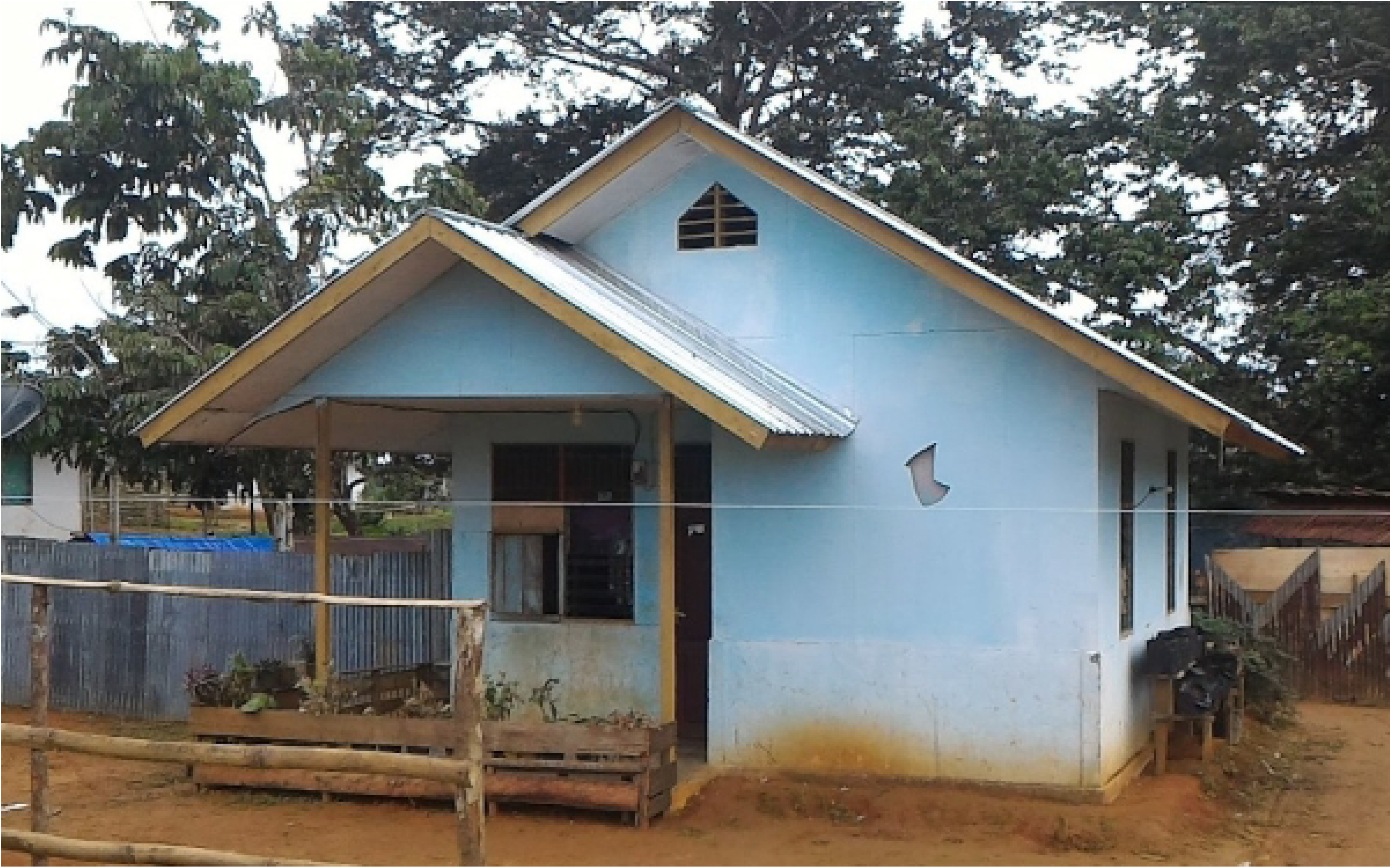
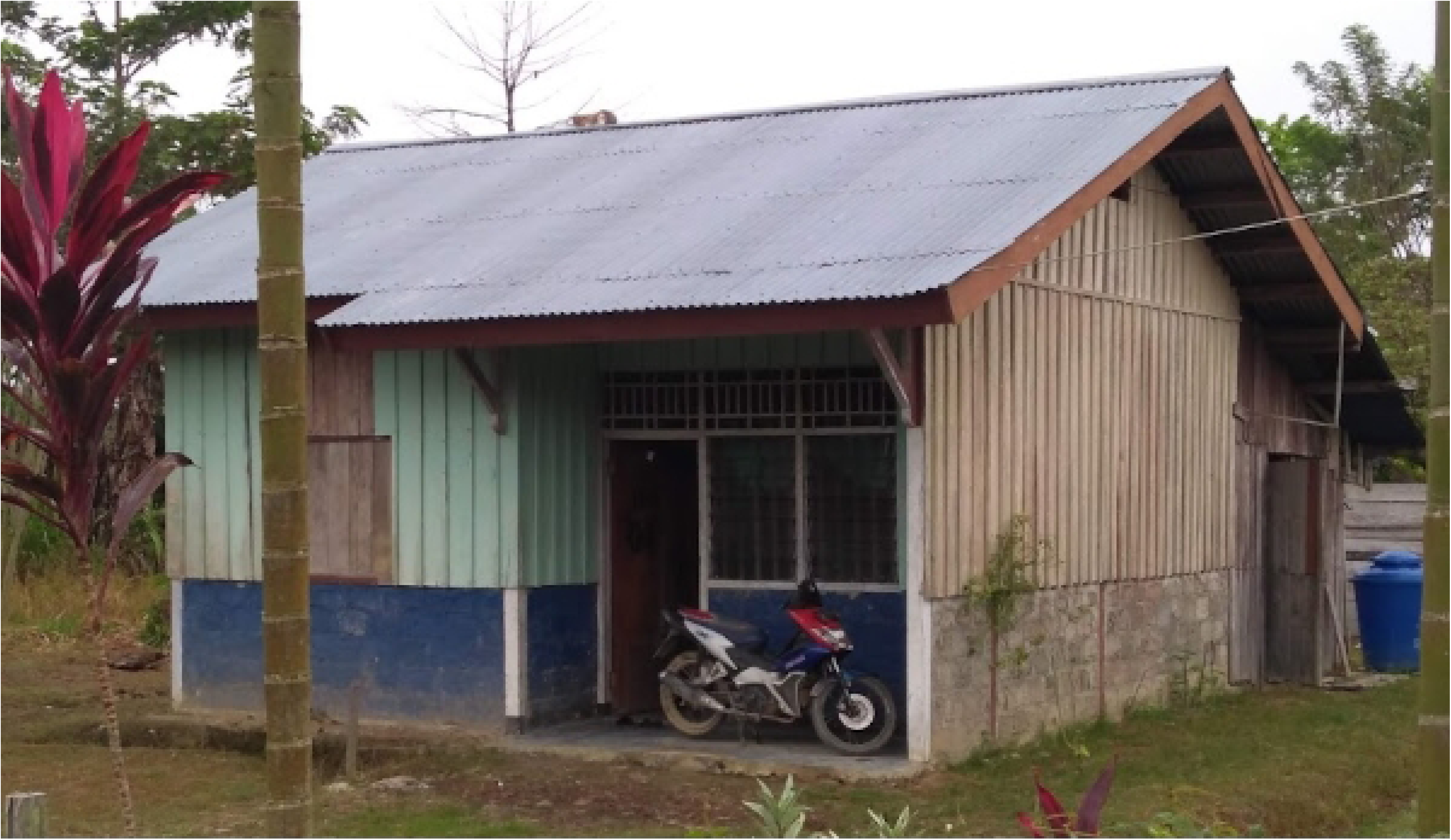
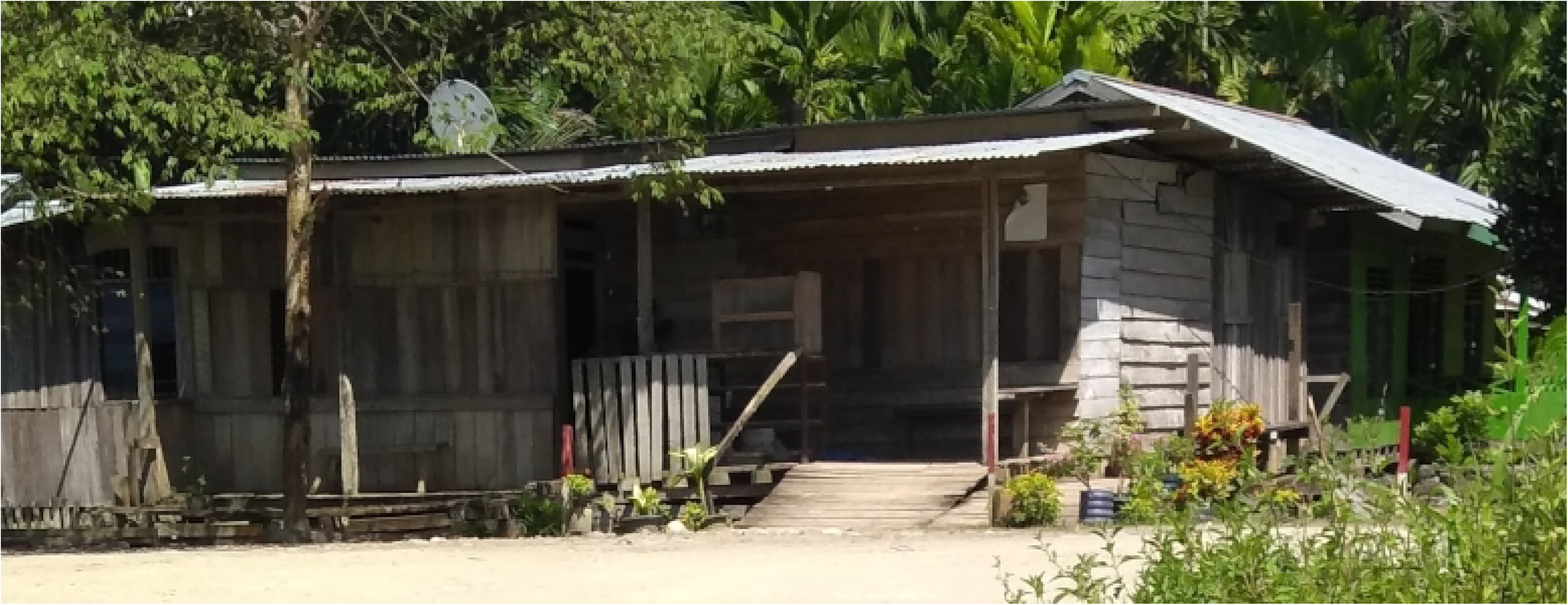
Type of houses in household survey. (a) and (b) full and half wooden wall with iron roof, (c) and (d) cement wall with iron roof.

Notably, in Asmat Regency, 96.2% of the houses were made from wooden planks, wooden blocks, and bamboo (Fig 3a). These homes were typically elevated more than 50 cm above the ground along riversides or river mouths, making them susceptible to tides and temporary water pooling. In contrast, 39% of houses in Yapen Islands, 45% in Mimika, and 61% in Sarmi were also made from wooden planks, but only 41% in Mimika and Sarmi and 22% in Yapen Islands were elevated more than 50 cm above the ground.

A clustering analysis based on wall and floor materials revealed four main house types. The largest category, 43%, consisted of houses made entirely from wooden planks for both walls and floors. The second largest category, 29.5%, included houses with cement or concrete walls and cement or ceramic floors. The third type, 16.9%, featured walls made from a mix of cement and wood or stone, with floors of cement or ceramic. The remaining 10.4% of houses had wooden plank walls and floors made from cement, ceramic, or dirt. On average, houses had 2.4 (±0.8) doors and 4.8 (±2.5) windows.

The majority of houses lacked mosquito-proofing features such as screened windows or doors. While 88.8% of houses had eaves, only about 25% (ranging from 15.0% to 35.4%) used mosquito nets to block eaves. Mosquito screens on doors, windows, and vents were observed across all four house types. Electricity was available in 91.3% of the surveyed houses, with 73.9% connected to the State Electricity Company (Perusahaan Listrik Nasional, PLN), and 12.6% and 13.5% powered by solar cells and generators, respectively.

Approximately 80.0% of interviewees reported that at least one member of their household had experienced a malaria infection (Table S3-3), with the highest incidence reported in Sarmi and Mimika (94.9% and 92.3%, respectively). Additionally, 25.5% of those interviewed mentioned having personally suffered from malaria. The prevalence of diagnosed malaria infections within the two weeks preceding the interview, or ongoing at the time of the interview, was also significant across the regions, averaging 13.1% and ranging from 5.8% to 20.3%. Furthermore, the average rate of malaria-related deaths within the last two years among surveyed households was 2.1%, with a range from 0.6% to 5.7%.

### Community practices for preventing mosquito bites

Based on the household survey results, malaria vector control interventions, particularly targeting adult mosquitoes through IRS spraying and ITN distribution, were implemented across the eight regencies (Fig 4 and Table S3-4). On average, IRS spraying covered 28.4% of the study areas, but it was not uniformly or simultaneously conducted in all locations. For example, in Keerom, 67.3% of the area was sprayed, with some villages fully covered, others partially, and one not sprayed at all. Over 60% of households in Jayapura, Mimika, and Yapen Islands that received IRS had been sprayed within three months prior to the survey. In contrast, in Keerom, Sarmi, and Waropen, a significant portion of the spraying occurred more than six months earlier. In Boven Digoel and Asmat, IRS was carried out 3–6 months and over six months before the survey, with similar percentages across both time frames. Most of the IRS activities had been conducted by health workers.

**Fig 4.**
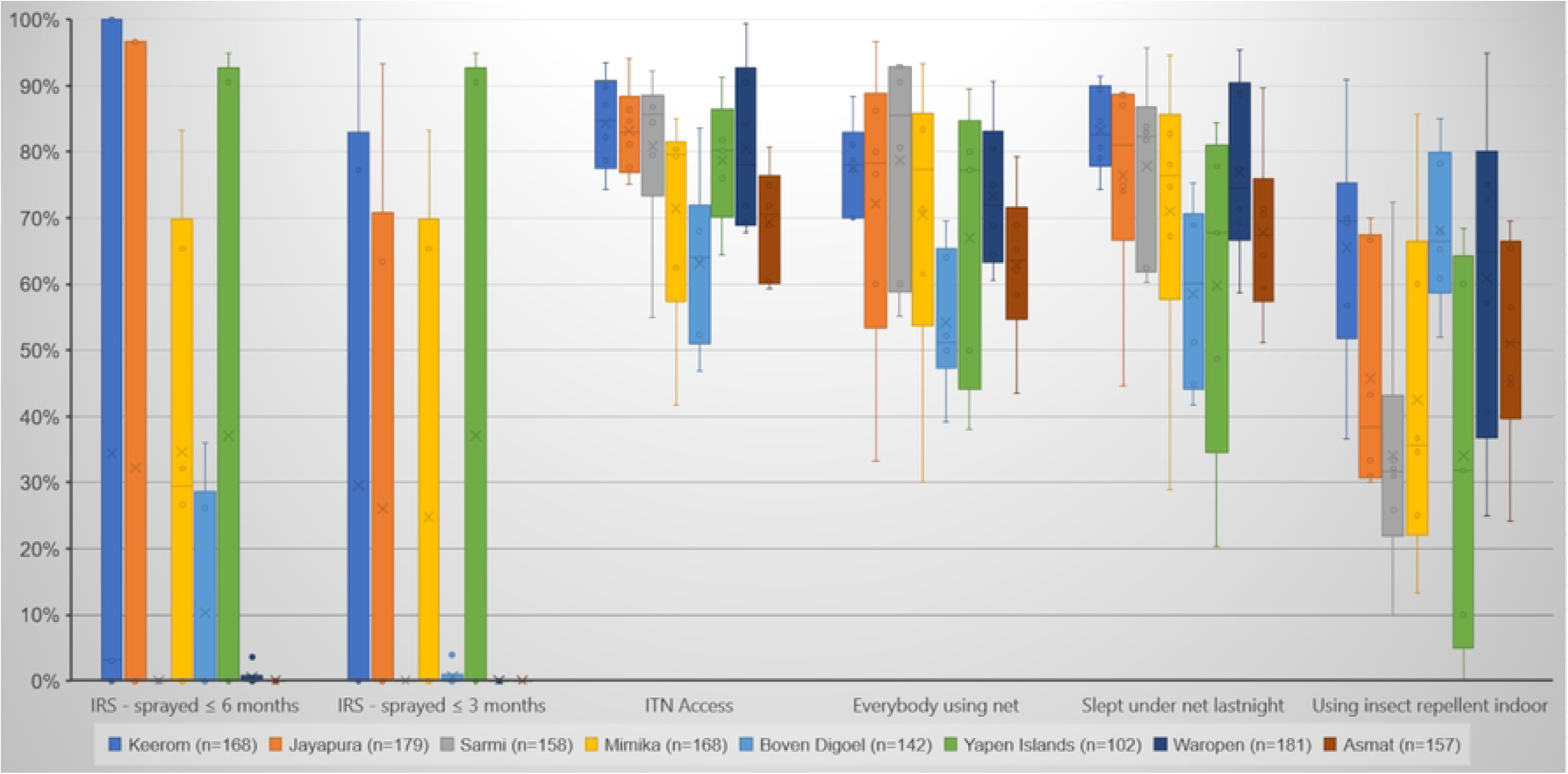
Mosquito bites prevention efforts.

Table 2 shows the estimated IRS effectiveness for potential protection in 14 villages that have mentioned in Table 1. Overall IRS effectiveness ranged from zero to no more than 25%, meaning that more than 75% of the *Anopheles* in these villages did not rest indoor, even though after took blood feeding indoor. This overall *Anopheles* behavior reflected similarly to the *An. koliensis* IRS effectiveness. Moreover, *An. punctulatus* mostly had less IRS effectiveness than *An. koliensis*, but in Konderjen and Mwarei the IRS effectiveness were 66.7% and 50.0% respectively. Meanwhile, *An. farauti* that tended to bite outdoors in the early evening [11,29], also had less IRS effectiveness, except in Sunum/Yamna village (36.4%).

**Table 2.** The estimated IRS effectiveness for potential protection in 14 villages.

| Village | Skofro | Pikere | Hanggey Among | Sunum / Yamna | Samanente | Konderjan | Arbais | Webrau | Mwarei | Kaugapu | Wanagon | Sukanggo | Migiwia | Yasiuw | All Villages |
| --- | --- | --- | --- | --- | --- | --- | --- | --- | --- | --- | --- | --- | --- | --- | --- |
| **Total *Anopheles*** |  |  |  |  |  |  |  |  |  |  |  |  |  |  |  |
| *Anopheles* indoor per house | 50.0 | 42.3 | 49.3 | 19.0 | 4.3 | 11.8 | 12.0 | 23.0 | 14.3 | 26.0 | 2.8 | 7.5 | 4.8 | 1.8 | 6.1 |
| *Anopheles* outdoor per house | 66.0 | 37.0 | 84.5 | 41.0 | 18.8 | 7.3 | 30.0 | 27.0 | 62.3 | 30.8 | 23.0 | 13.0 | 9.8 | 12.3 | 10.8 |
| *Anopheles* per house | 116.0 | 79.3 | 133.8 | 60.0 | 23.0 | 19.0 | 42.0 | 50.0 | 76.5 | 56.8 | 25.8 | 20.5 | 14.5 | 14.0 | 16.9 |
| IRD for total *Anopheles* | 6.3 | 0.0 | 14.0 | 12.8 | 1.5 | 3.5 | 8.8 | 10.3 | 17.0 | 5.0 | 0.0 | 2.3 | 0.3 | 1.0 | 2.1 |
| Potential of IRS | 5.4% | 0.0% | 10.5% | 21.3% | 6.5% | 18.4% | 20.8% | 20.5% | 22.2% | 8.8% | 0.0% | 11.0% | 1.7% | 7.1% | 12.5% |
| **Total *An. koliensis*** |  |  |  |  |  |  |  |  |  |  |  |  |  |  |  |
| *An. koliensis* indoor per house | 38.3 | 16.0 | 39.3 | 14.3 | 2.3 | 11.0 | 11.5 | 18.0 | 12.5 | 21.8 | 2.3 | 4.3 | 0.3 | 1.8 | 4.3 |
| *An. koliensis* outdoor per house | 49.0 | 12.3 | 73.0 | 32.8 | 11.8 | 6.0 | 24.5 | 22.0 | 51.3 | 25.5 | 18.3 | 9.8 | 0.3 | 11.8 | 8.0 |
| *An. koliensis* per house | 87.3 | 28.3 | 112.3 | 47.0 | 14.0 | 17.0 | 36.0 | 40.0 | 63.8 | 47.3 | 20.5 | 14.0 | 0.5 | 13.5 | 12.3 |
| IRD for *An. koliensis* | 5.0 | 0.0 | 12.3 | 10.8 | 1.0 | 3.0 | 8.3 | 8.5 | 13.0 | 4.8 | 0.0 | 1.3 | 0.0 | 1.0 | 1.5 |
| Potential of IRS | 5.7% | 0.0% | 10.9% | 22.9% | 7.1% | 17.6% | 22.9% | 21.3% | 20.4% | 10.1% | 0.0% | 8.9% | 0.0% | 7.4% | 12.6% |
| **Total *An. punctulatus*** |  |  |  |  |  |  |  |  |  |  |  |  |  |  |  |
| *An. punctulatus* indoor per house | 10.8 | 24.0 | 9.5 | 3.3 | 1.8 | 0.5 | 0.5 | 3.5 | 0.5 | 2.5 | 0.3 | 2.0 | 0.0 | 0.0 | 1.3 |
| *An. punctulatus* outdoor per house | 13.0 | 22.3 | 11.0 | 6.3 | 6.0 | 0.3 | 5.3 | 3.5 | 3.0 | 1.8 | 1.3 | 0.5 | 0.0 | 0.0 | 1.7 |
| *An. punctulatus* per house | 23.8 | 46.3 | 20.5 | 9.5 | 7.8 | 0.8 | 5.8 | 7.0 | 3.5 | 4.3 | 1.5 | 2.5 | 0.0 | 0.0 | 3.0 |
| IRD for *An. punctulatus* | 0.8 | 0.0 | 1.8 | 1.0 | 0.0 | 0.5 | 0.5 | 1.0 | 1.8 | 0.0 | 0.0 | 0.3 | 0.0 | 0.0 | 0.2 |
| Potential of IRS | 3.2% | 0.0% | 8.5% | 10.5% | 0.0% | 66.7% | 8.7% | 14.3% | 50.0% | 0.0% | 0.0% | 10.0% | NA | NA | 5.5% |
| **Total *An. farauti*** |  |  |  |  |  |  |  |  |  |  |  |  |  |  |  |
| *An. farauti* indoor per house | 0.0 | 0.3 | 0.0 | 1.3 | 0.0 | 0.0 | 0.0 | 1.3 | 0.5 | 1.5 | 0.0 | 0.0 | 4.3 | 0.0 | 0.3 |
| *An. farauti* outdoor per house | 0.0 | 0.3 | 0.0 | 1.5 | 0.3 | 0.0 | 0.0 | 1.0 | 6.5 | 2.8 | 3.5 | 1.0 | 9.3 | 0.5 | 0.7 |
| *An. farauti* per house | 0.0 | 0.5 | 0.0 | 2.8 | 0.3 | 0.0 | 0.0 | 2.3 | 7.0 | 4.3 | 3.5 | 1.0 | 13.5 | 0.5 | 1.0 |
| IRD for *An. farauti* | 0.0 | 0.0 | 0.0 | 1.0 | 0.0 | 0.0 | 0.0 | 0.3 | 1.0 | 0.3 | 0.0 | 0.0 | 0.3 | 0.0 | 0.1 |
| Potential of IRS | NA | 0.0% | NA | 36.4% | 0.0% | NA | NA | 11.1% | 14.3% | 5.9% | 0.0% | 0.0% | 1.9% | 0.0% | 7.9% |

Note: NA in potential of IRS means “not applicable”, due to the calculation was divided by zero.

The interview results regarding implementing insecticide-treated nets (ITNs), as presented in Fig 4 and Table S3-4, indicate that ITNs were generally distributed evenly and used appropriately. The average ITN coverage closely matched the number of bedrooms per house, suggesting that nearly every bedroom could have an ITN installed. Additionally, the average ITN access was 75.8% (ranging from 63.1% to 84.0%), meaning that three out of four people had access to an ITN, with one ITN typically shared by up to two people.

The proportion of household members using ITNs was quite high, with 70.3% of all individuals in the household (including adult males, adult females, and children) reported to be using ITNs. Furthermore, 72.7% of household members (an average of 3.9 persons per household) slept under an ITN the night before the survey. However, reasons for not using ITNs varied, with the most common being discomfort due to heat inside the net. Other frequently cited reasons included the belief that there was no need to use ITNs because the area had already been sprayed with mosquito repellent. Approximately half of the surveyed households used indoor insect repellents, with mosquito coils and sprays being the most common methods to prevent mosquito bites.

### Nighttime habits and usage of mosquito repellent

Questions about nighttime activities were asked to assess the risk of exposure to mosquito bites, with the results presented in Fig 5 and Table S3-5. The majority of evening meal activities (77.1%) occurred between 19:00 and 21:00, and most people ate at home. However, on average, 3.3% of interviewees reported having their evening meal outside the house. Additionally, 21.9% of respondents mentioned resting outside the house after eating.

**Fig 5.**
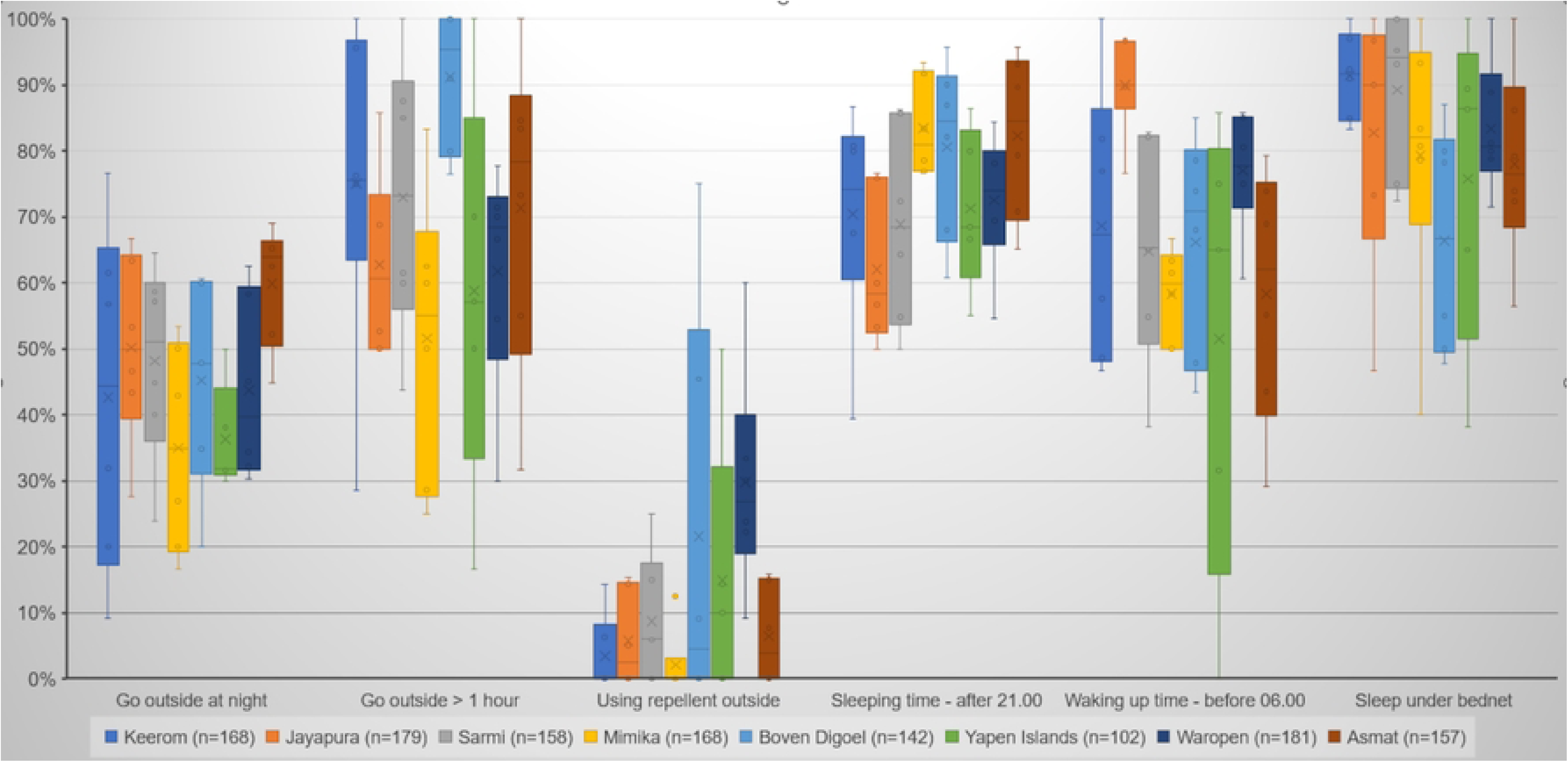
Human behavior during night time.

Outdoor nighttime activities were common, with 46.1% of interviewees engaging in such activities, and the percentage was fairly consistent across the study sites, ranging from 36.3% to 59.9%. These activities primarily included sitting around in the house yard or visiting friends and relatives in the neighborhood (71.6%) and going fishing or hunting (16.3%). Among those who engaged in nighttime outdoor activities, 68.3% reported that these activities lasted for more than an hour. However, only 10.9% of interviewees used mosquito repellent while outside at night.

Most respondents (73.4%) reported going to sleep after 21:00, and 67.5% woke up before 06:00. A significant majority (81.6%) stated that they slept under an ITN. Only a small percentage of interviewees (0.4%) reported sleeping outside the house.

### Gaps in protection

Table 3 presents gaps in protection identified from the household survey and examines their correlations with calculated house types and household socioeconomic status in the study area. The house mosquito screen and IRS coverage were included as major gaps in protection, meanwhile ITN coverage and usage was included as minor to moderate gaps. Moreover, although nighttime activities that exposed to mosquito bites were not included as major gaps in protection, because of resided in the malaria endemic area these activities could increase malaria transmission. The analysis correlations revealed that, with p-values greater than 0.05, most gaps in protection did not show a significant correlation with either house types or household socioeconomic status.

**Table 3.** Gaps in protection and correlations with house type and socioeconomic status.

| Gaps in protection |  | House Type |  |  |  | Chi-Square Test (p-value) | Socioeconomic status (Wealth Index) |  |  |  |  | Chi-Square Test (p-value) |
| --- | --- | --- | --- | --- | --- | --- | --- | --- | --- | --- | --- | --- |
|  |  | n = 542 | n = 212 | n = 370 | n = 131 |  | n = 323 | n = 524 | n = 175 | n = 179 | n = 54 |  |
|  |  | type one (%) | type two (%) | type three (%) | type four (%) |  | 1<sup>st</sup> quintile (%) | 2<sup>nd</sup> quintile (%) | 3<sup>rd</sup> quintile (%) | 4<sup>th</sup> quintile (%) | 5<sup>th</sup> quintile (%) |  |
| Protection from mosquitoes entering house |  |  |  |  |  |  |  |  |  |  |  |  |
| Screened door | +++ | 24 (1.9%) | 9 (0.7%) | 15 (1.2%) | 4 (0.3%) | 0.9155 | 11 (0.9%) | 25 (2.0%) | 10 (0.8%) | 6 (0.5%) | 0 (0.0%) | 0.3233 |
| Screened window | +++ | 30 (2.4%) | 12 (1.0%) | 28 (2.2%) | 5 (0.4%) | 0.3947 | 16 (1.3%) | 31 (2.5%) | 12 (1.0%) | 14 (1.1%) | 2 (0.2%) | 0.6578 |
| Screened eave (n=1115) | +++ | 114 (10.2%) | 56 (5.0%) | 76 (6.8%) | 32 (2.9%) | 0.2993 | 67 (6.0%) | 120 (10.8%) | 39 (3.5%) | 48 (4.3%) | 4 (0.4%) | 0.03364 |
| Mosquito bites preventions |  |  |  |  |  |  |  |  |  |  |  |  |
| IRS |  |  |  |  |  |  |  |  |  |  |  |  |
| All house sprayed < 3 months | +++ | 74 (5.9%) | 22 (1.8%) | 54 (4.3%) | 15 (1.2%) | 0.4654 | 42 (3.3%) | 64 (5.1%) | 27 (2.2%) | 26 (2.1%) | 6 (0.5%) | 0.7913 |
| All house sprayed < 6 months | +++ | 102 (8.1%) | 30 (2.4%) | 73 (5.8%) | 20 (1.6%) | 0.2845 | 61 (4.9%) | 87 (6.9%) | 40 (3.2%) | 30 (2.4%) | 7 (0.6%) | 0.3099 |
| ITN access | + | 76.6% | 74.5% | 75.9% | 74.1% | - | 74.6% | 77.3% | 78.6% | 71.9% | 72.8% | - |
| Everybody using ITNs | ++ | 383 (30.5%) | 145 (11.6%) | 266 (21.2%) | 87 (6.9%) | 0.6171 | 225 (17.9%) | 365 (29.1%) | 130 (10.4%) | 118 (9.4%) | 43 (3.4%) | 0.2508 |
| ITN age (n=1001) |  |  |  |  |  |  |  |  |  |  |  |  |
| > 3 years | + | 32 (3.2%) | 14 (1.4%) | 28 (2.8%) | 6 (0.6%) | 0.4815 | 9 (0.9%) | 38 (3.8%) | 9 (0.9%) | 16 (1.6%) | 8 (0.8%) | 0.001005 |
| ITN conditions (n=999) |  |  |  |  |  |  |  |  |  |  |  |  |
| Lots of holes | + | 58 (5.8%) | 23 (0.0%) | 36 (0.0%) | 7 (0.0%) | 0.4164 | 29 (0.0%) | 55 (0.0%) | 18 (0.0%) | 17 (0.0%) | 5 (0.0%) | 0.9791 |
| NOT using indoor repellent | ++ | 263 (21.0%) | 105 (8.4%) | 190 (15.1%) | 66 (5.3%) | 0.8661 | 167 (13.3%) | 252 (20.1%) | 92 (7.3%) | 89 (7.1%) | 24 (1.9%) | 0.6928 |
| Outdoor activities at night |  |  |  |  |  |  |  |  |  |  |  |  |
| Going outside at night | ++ | 247 (19.7%) | 103 (8.2%) | 164 (13.1%) | 64 (5.1%) | 0.6923 | 146 (11.6%) | 239 (19.0%) | 84 (6.7%) | 84 (6.7%) | 25 (2.0%) | 0.9774 |
| Going outside at night (n=578) |  |  |  |  |  |  |  |  |  |  |  |  |
| Time progress > 1 hour | + | 159 (27.5%) | 74 (12.8%) | 115 (19.9%) | 50 (8.7%) | 0.1428 | 110 (19.0%) | 162 (28.0%) | 54 (9.3%) | 53 (9.2%) | 19 (3.3%) | 0.2222 |
| NOT using repellent outside | +++ | 216<br>(37.4%) | 91<br>(15.7%) | 150<br>(26.0%) | 58<br>(10.0%) | 0.6043 | 134<br>(23.2%) | 213<br>(36.9%) | 76<br>(13.1%) | 71<br>(12.3%) | 21 (3.6%) | 0.4445 |
| **Sleeping behavior** |  |  |  |  |  |  |  |  |  |  |  |  |
| Sleeping time |  |  |  |  |  |  |  |  |  |  |  |  |
| After 21.00 | ++ | 397<br>(31.6%) | 167<br>(13.3%) | 267<br>(21.3%) | 95 (7.6%) | 0.3312 | 245<br>(19.5%) | 390<br>(31.1%) | 137<br>(10.9%) | 119<br>(9.5%) | 35 (2.8%) | 0.04318 |
| Waking up time |  |  |  |  |  |  |  |  |  |  |  |  |
| Before 06.00 | ++ | 366<br>(29.2%) | 144<br>(11.5%) | 249<br>(19.8%) | 91 (7.3%) | 0.9733 | 220<br>(17.5%) | 358<br>(28.5%) | 118<br>(9.4%) | 124<br>(9.9%) | 30 (2.4%) | 0.4102 |
| Sleep outdoor | + | 3 (0.2%) | 2 (0.2%) | 1 (0.1%) | 0 (0.0%) | 0.5735 | 1 (0.1%) | 4 (0.3%) | 0 (0.0%) | 1 (0.1%) | 0 (0.0%) | 0.6966 |
| Sleep NOT under ITNs | + | 89 (7.1%) | 42 (3.3%) | 68 (5.4%) | 31 (2.5%) | 0.2486 | 73 (5.8%) | 92 (7.3%) | 25 (2.0%) | 35 (2.8%) | 5 (0.4%) | 0.05619 |

Note:
Gaps in protection column: “+”, “++” and “+++” mean gaps in protection with different magnitude of the problems, i.e. minor, moderate and major respectively.

## DISCUSSION

This study highlights several gaps in protection [9] against mosquito bites, both indoors and outdoors, which may contribute to the recent stagnation in reducing malaria incidence in Papua. To achieve malaria elimination by 2030, decision-makers in Papua Province must accelerate efforts to reduce malaria cases to zero. This will require addressing both indoor and outdoor transmission through enhanced and targeted mitigation strategies targeting both vectors and humans. The utilization of multiple types of data (human behavior and entomology) enables a direct understanding of where and when transmission events occur – outputs that enable the mapping of specific interventions and how they function onto these spatial and temporal gaps in protection.

To effectively combat malaria transmission, both indoor and outdoor gaps in mosquito bite protection must be addressed simultaneously. This study identified the Punctulatus group as the primary malaria vector across Papua, which bites humans both indoors and outdoors with equal frequency [11,30]. The current malaria control program by the Ministry of Health focuses on early diagnosis and treatment, ITN distribution, and indoor residual spraying (IRS) [1,2]. However, this approach predominantly targets indoor transmission and does not address the outdoor transmission component.

To enhance indoor protection, identified gaps such as the suboptimal use of ITNs, the absence of house mosquito screens [31–33], and insufficient IRS coverage need to be addressed. Improving these measures will optimize indoor protection during nighttime.

For mitigating outdoor transmission, it may be crucial to reduce larval habitats, particularly those near residential areas, through larval source management [11,34]. This approach has the potential to significantly decrease the local *Anopheles* population, thereby reducing overall malaria transmission. Additionally, integrating health education (Social and Behavior Change Communication (SBC)) for local inhabitants to minimize mosquito exposure during nighttime activities is recommended [10], and may increase ITN use. Human behavior observations (HBO) conducted during HLC in Papua [15,18,35–37] have highlighted these gaps in the protection and local drivers of malaria transmission.

The distribution of ITNs across the eight regencies in this study demonstrated nearly complete coverage, with each household having approximately three ITNs. Despite this, the household survey revealed that while over 70% of household members used ITNs and more than 72% slept under them the night before the interview, gaps in protection remain due to human behavior.

Even with widespread ITN coverage, the issue of people being indoors at night without bed net protection persists. To address this, options include regular IRS or the installation of mosquito screens on doors and ventilation openings. Additionally, innovative vector control tools like spatial repellents could be used. These tools offer protection for indoor spaces and have been shown to effectively reduce malaria transmission in various field settings [38–40].

The habit of people in Papua engaging in nighttime outdoor activities significantly contributes to malaria infection [9,10,15,41,42], irrespective of house type or socioeconomic status. Survey results indicate that these activities often take place in the neighborhood, such as visiting friends or attending cultural and religious events. Nighttime activities like hunting and fishing pose an even higher risk than mere socializing in the neighborhood. To address this protection gap, health education on malaria transmission and the use of household insecticides and repellents is recommended. Educating the community about the risks associated with outdoor activities and promoting protective measures can help reduce exposure to mosquito bites and, consequently, malaria transmission.

### Limitations of the study

Given that this study was a cross-sectional survey conducted over a short period of time, several limitations should be noted. A larger sampling frame would have provided more representative data. Sampling at monthly intervals throughout the year would have offered a more comprehensive understanding of vector and human behaviors across different seasons and variations in human activities. Additionally, the data collected from selected sentinel sites may not fully represent the entire regency, potentially affecting the generalizability of the findings. Furthermore, missing data from both the HBO and household questionnaire datasets may have influenced the accuracy and reliability of site-specific analyses.

## CONCLUSIONS

The evidence underscores the shortcomings of existing malaria control efforts in Papua Province and emphasizes the need to address both indoor and outdoor mosquito bite protection gaps. To improve protection, it is recommended to enhance indoor measures while also implementing strategies to reduce outdoor transmission, such as community-driven larval source management to eliminate mosquito breeding sites near dwellings. Novel strategies such as spatial repellents may also be evaluated in scaled studies. In conclusion, this study identified several gaps in protection against mosquito bites both indoors and outdoors across eight regencies in Papua Province. Key issues include inadequate ITN usage, insufficient IRS coverage, limited indoor protection before sleeping, absence of house screening, and the habit of engaging in outdoor activities without protection. These findings highlight the need to optimize indoor protection measures to effectively mitigate indoor malaria transmission. However, current malaria control activities have not sufficiently addressed outdoor mosquito exposure.

## Data Availability

All datasets generated and analyzed during this study are included in the manuscript

## ACKNOWLEDGEMENTS

We extend our sincere appreciation to UNICEF for supporting this activity in Papua. Our heartfelt thanks go to the laboratory and field study teams for their efforts in sample and data collection. We are also deeply grateful to the Eijkman Research Center for Molecular Biology and the National Research and Innovation Agency (BRIN) for their ongoing support. We express our profound gratitude to the residents in the study area for their time, patience, and participation in this study. Special thanks are extended to the Health Departments of the Keerom, Boven Digul, Mimika, Sarmi, Jayapura, Yapen Islands, Waropen, and Asmat regencies, as well as the Primary Health Departments, for their invaluable support. We also acknowledge the dedication of the entomology team, local field workers, data entry clerks, and the numerous local volunteers who contributed tirelessly to ensure the quality of the data and the success of this project.

## Author Contribution

IER, LS, DHP, PBSA, NFL, and DS were responsible for the study design, supervised the data collection, and contributed to the first draft writing of the manuscript. IER, LS, DHP, PBSA, MES, NFL, WAH, and DS performed the data collection, laboratory work, and analysis. IER, PBSA, NFL, and DS drafted the manuscript. All authors are main contributors who have read and approved the final manuscript.

## Availability of data and materials

All datasets generated and analyzed during this study are included in the manuscript.

## Declarations

### Consent for publication

Not applicable.

## SUPPORTING INFORMATION

**S1 File Part A. Human behavior observation (HBO).**

**S1 File Part B. Household survey.**

**S2 File. Human behavior observations and adjusted-human biting rates in 14 villages.**

**S3 File. Household survey results.**

